# Knowledge, attitudes, and perceptions of healthcare students and professionals on the use of artificial intelligence in healthcare

**DOI:** 10.1101/2024.01.08.24300977

**Authors:** Muhammad Mustafa Habib, Zahra Hoodbhoy, M. A. Rehman Siddiqui

**Affiliations:** The Aga Khan University, Karachi, Pakistan; Department of Pediatrics and Child Health, The Aga Khan University, Karachi, Pakistan; Department of Ophthalmology and Visual Sciences, The Aga Khan University Hospital, Karachi, Pakistan

**Keywords:** Artificial Intelligence, Healthcare professionals, Pakistan, Knowledge, Attitudes, and Perceptions, Machine learning, Big data

## Abstract

The emergence of artificial intelligence (AI) technologies has emerged as a promising solution to enhance healthcare efficiency and improve patient outcomes. The objective of this study is to analyse the knowledge, attitudes, and perceptions of healthcare professionals in Pakistan about AI in healthcare.

We conducted a cross-sectional study using a questionnaire distributed via Google Forms. This was distributed to healthcare professionals (e.g., doctors, nurses, medical students, and allied healthcare workers) working or studying in Pakistan. The questions were related to participant demographics, basic understanding of AI, AI in education and practice, AI applications in healthcare systems, AI’s impact on healthcare professions and the socio-ethical consequences of the use of AI. We analyzed the data using Statistical Package for Social Sciences (SPSS) statistical software, version 26.0.

Overall, 616 individuals responded to the survey while n=610 (99.0%) of respondents consented to participate. The mean age of participants was 32.2 ± 12.5 years. Most of the participants (78.7%, n=480) had never received any formal sessions or training in AI during their studies/employment. A majority of participants, 70.3% (n=429), believed that AI would raise more ethical challenges in healthcare. In all, 66.4% (n=405) of participants believed that AI should be taught at the undergraduate level.

The survey suggests that there is insufficient training about AI in healthcare in Pakistan despite the interest of many in this area. Future work in developing a tailored curriculum regarding AI in healthcare will help bridge the gap between the interest in use of AI and training.

**Author summary:** In our research article titled, “Knowledge, Attitudes, and Perceptions of Healthcare Students and Professionals on the Use of Artificial Intelligence in Healthcare,” we set out to explore the scope of AI understanding within the healthcare community in Pakistan. We were particularly motivated to bridge the existing gaps in knowledge and explore the once uncharted territories of AI perception among all healthcare staff, including nurses, medical students, and allied healthcare workers.

Our study is particular;y significanct as it goes beyond the traditional investigations previously conducted in Pakistan, incorporating a holistic approach to assess opinions across all healthcare roles. By doing so, we aimed to provide a complete understanding of AI’s impact and potential in a developing country like Pakistan. Our findings shed light on these previously unexplored perspectives and help in contributing valuable insights for both local healthcare professionals and the broader global community.

Pakistan is a region with unique challenges - this research aims to serve as a foundation for future discussions on the integration of AI in healthcare. By providing a comprehensive view of attitudes and perceptions, we aim to foster informed discussions and strategic planning for the effective utilization and adoption of AI and ultimately, enhancing healthcare systems, efficiency, and delivery.

## Introduction

The field of medicine and healthcare stands on the precipice of a revolutionary transformation due to the immense potential of artificial intelligence (AI). AI has a myriad of healthcare applications – ranging from improving diagnostic accuracy, predicting patient outcomes, and even providing personalized treatment plans. One such instance is found in the field of radiology. Here, AI can utilize advanced deep learning techniques to enable the classification of chest radiographs based on abnormalities and facilitating triage of patients [2]. Additionally, AI continues to play an integral role in shaping the way medical students and future healthcare professionals interact within the healthcare ecosystem. Evidence exists that AI solutions offer a new horizon of possibilities for learning and higher education, particularly for medical and nursing students [3]. Furthermore, research suggests that AI can help nurses assume an even greater role in healthcare delivery by offering sophisticated tools to support nurses anytime/anywhere enabling nurses to fulfil a practitioner role and delivering care across the continuum [4].

AI integration in healthcare and education has seen widespread acceptance in high-income countries (HICs), as evidenced by a 2020 survey of medical students in the United Kingdom, which revealed generally positive attitudes and perceptions toward AI’s incorporation into medical curricula. A majority (88%, n = 432) of respondents from this study believed that AI would play an important role in healthcare in the future. Surprisingly, only 9.2% (n=45) of students from this survey received some form of teaching on AI [5]. Interestingly, this was similar to a study from Pakistan where only 33 respondents (9.9%) had obtained some form of training in AI during their education [6]. It is important to note that the utilization of AI in healthcare remains relatively unexplored, under-researched, and underfunded, particularly in low-and middle-income countries (LMICs) like Pakistan [7]. Existing data from previous studies from Pakistan indicates a growing interest among medical students in AI, even in the absence of specialized training [6][8], which presents a noteworthy trend to be explored further. This is particularly interesting in the context of Pakistan, as AI, if leveraged appropriately, holds tremendous promise for transforming the provision of healthcare services in resource-poor settings [9].

It is worth noting that prior studies have left many important questions unanswered. One significant gap in existing literature is the lack of data from other crucial segments of healthcare professionals, such as nurses and allied healthcare practitioners. For the successful integration of AI into Pakistan’s healthcare landscape, it is essential to establish a comprehensive and holistic understanding of all participants within the healthcare ecosystem. Additionally, the study aims to provide a comparative analysis with global literature and trends and guide policy and decision-making to facilitate the responsible and effective use of AI in Pakistani healthcare and build on existing literature and address the lack of data available from comparable LMICs and healthcare professionals besides doctors and medical students.

## Results

In total, 616 healthcare professionals initiated the survey, and 99.03% (n=610) of those who started the survey completed it. Among the respondents of this survey, the majority were males (n=357, 58.5%). The mean age of all participants was 32.23 ± 12.45 years ( Table 1).

**Table 1.** Participant sociodemographic characteristics.

| Demographic Question | Item | *n=610* | % | Mean | SD |
| --- | --- | --- | --- | --- | --- |
| Age (Years) |  |  |  | 32.23 | 12.45 |
| Gender | Female | 251 | 41.10 |  |  |
|  | Male | 357 | 58.50 |  |  |
|  | Prefer not to say | 2 | 0.30 |  |  |
| Highest educational qualification | Doctoral Degree | 97 | 15.90 |  |  |
|  | Master's degree | 133 | 21.80 |  |  |
|  | Associate Ordinary Bachelor | 4 | 0.70 |  |  |
|  | Bachelor's Degree | 325 | 53.30 |  |  |
|  | Matriculation level or below | 4 | 0.70 |  |  |
|  | Higher Secondary School Certificate | 40 | 6.60 |  |  |
|  | Secondary school Certificate | 7 | 1.10 |  |  |
| Role in healthcare setup | Doctor | 239 | 39.20 |  |  |
|  | Nurse | 43 | 7.00 |  |  |
|  | Allied Health Professional | 85 | 13.90 |  |  |
|  | Undergraduate student | 198 | 32.50 |  |  |
|  | Other | 45 | 7.40 |  |  |
| What is the classification of your place of work/studies? | Government | 388 | 63.60 |  |  |
|  | Private | 175 | 28.69 |  |  |
|  | Other | 47 | 7.71 |  |  |

### Participant demographics

Among our participants, 166 (44.7%) participants were enrolled in an MBBS program, while 50 (13.5%) were in BScN and PGME programs (16.4%, n=61). Overall, 307 (82.7%) of the participants had worked or were working in a clinical setting. Most of the respondents were either employed or studying at a private healthcare institute (64.1%, n=388), while only 28.9% (n=175) belonged to government healthcare institutions. The remaining 8 % (n=47) belonged to setups with a variety of designations, e.g., Trust Hospitals, NGOs, or military setups.

### Basic understanding of AI

A third of the participants, (30.7%; n=187) strongly or somewhat agreed that they were technologically adept, while only 10.4% (n=62) disagreed. Participants were then asked a series of questions to determine their general understanding of AI and AI-specific tools and applications (Fig 1).

**Fig 1.**
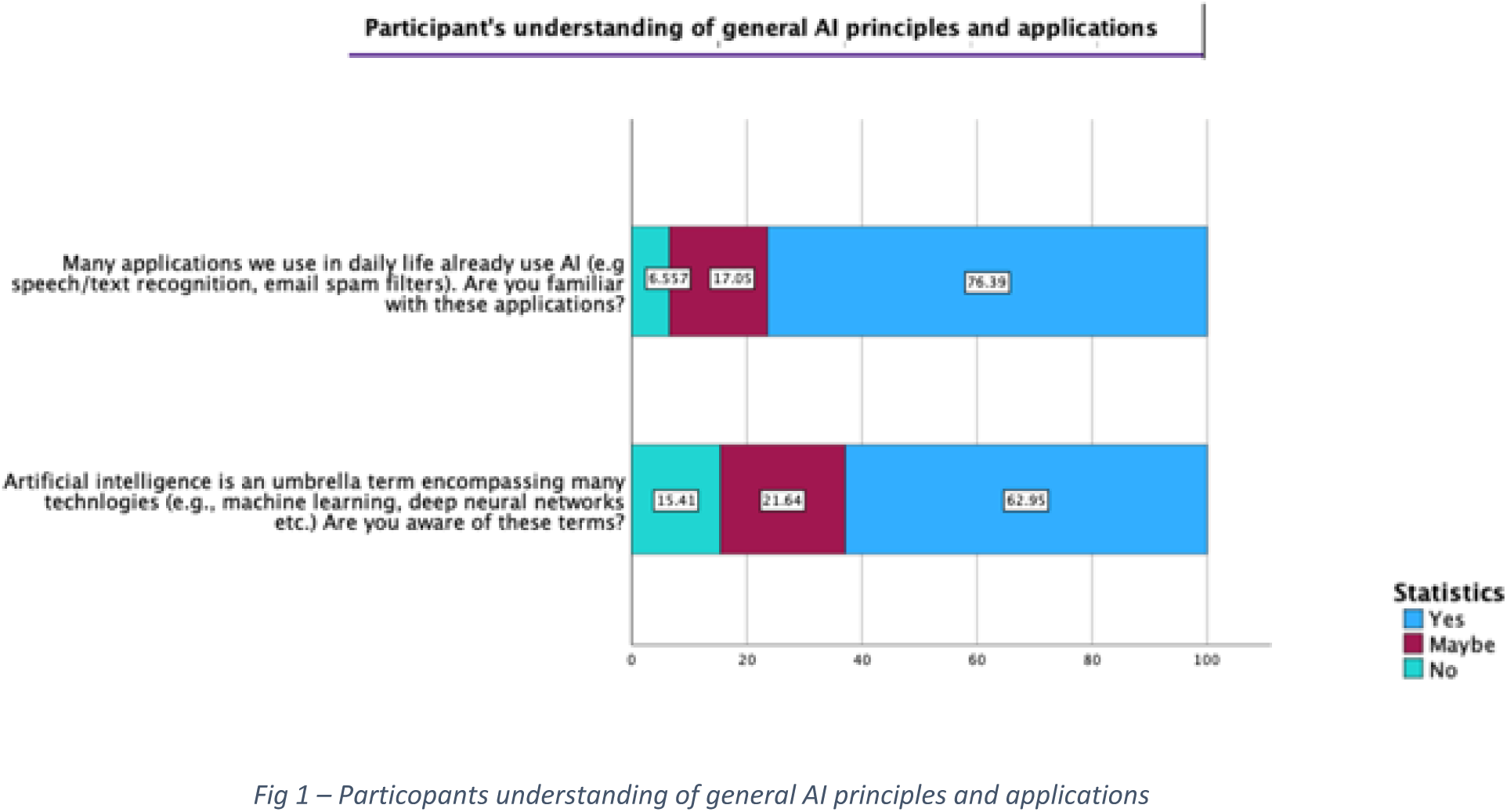
Particopants understanding of general AI principles and applications.

### AI in education

Most of the participants (78.7%, n=480) had never had any formal sessions or training in AI during their studies/employment. However, 254 (41.6%) participants strongly agreed or somewhat agreed (35.4%, n=216) that AI should be part of a healthcare professional’s training. Of the 610 respondents, 66.4% (n=405) believed that it would be essential for training in AI competencies to begin at the undergraduate level for students to prepare them for clinical practice.

The top three choices for sources of information about AI among the respondents were social media (66.4%, n=151), followed by web-browsing (48%, n=293), and information obtained from their school or workplace (31.3%, n=191). Respondents were also asked what the best methods would be to learn about AI in healthcare. The most popular methods identified by participants included workshops on programming languages in AI (67.4%, n=411), AI symposiums with experts (53.4%, n=326), and student-led journal clubs (25.7%, n=157).

Furthermore, participants were asked to comment on how much time they would like to spend learning about AI every month. A majority of participants (29%, n=177) wished to invest 1-5 hours every month while only a minority (5.9% n=36) were not interested in learning about AI (Fig 2).

**Fig 2.**
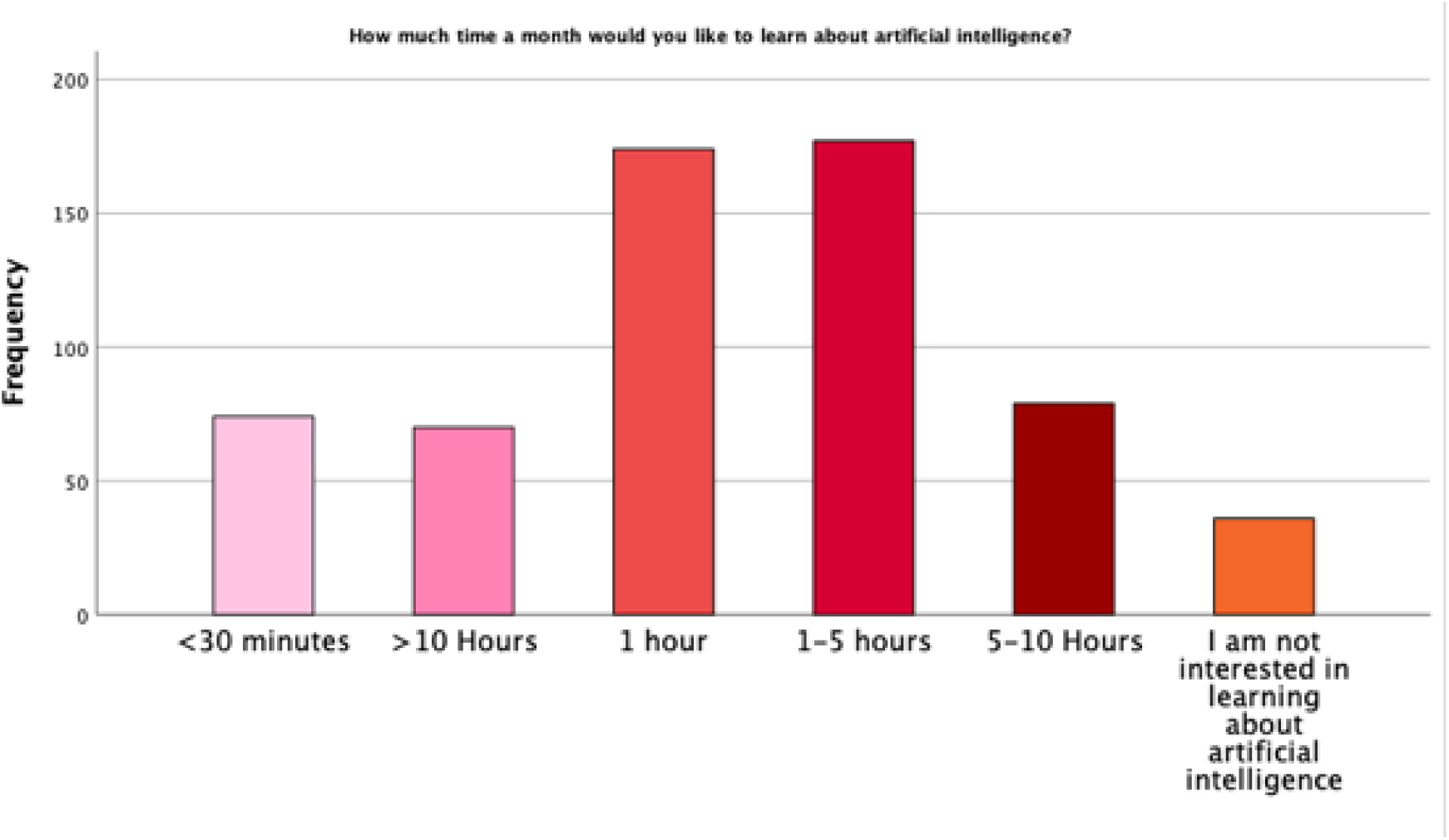
Participants were asked how much time they would like to spend learning about AI every month.

### AI Capabilities in healthcare systems

Participants were asked whether they believed that AI could perform certain tasks in a manner comparable to a human healthcare professional (Fig 3). Our findings suggested that participants were more likely than not to agree with statements regarding AI capabilities with regards to treatment planning, monitoring, diagnostic interpretation and analysing patient information. A majority of participants (56.1%, n=342) believed that it was either extremely unlikely or unlikely that AI would be able to provide empathetic care to patients.

**Fig 3.**
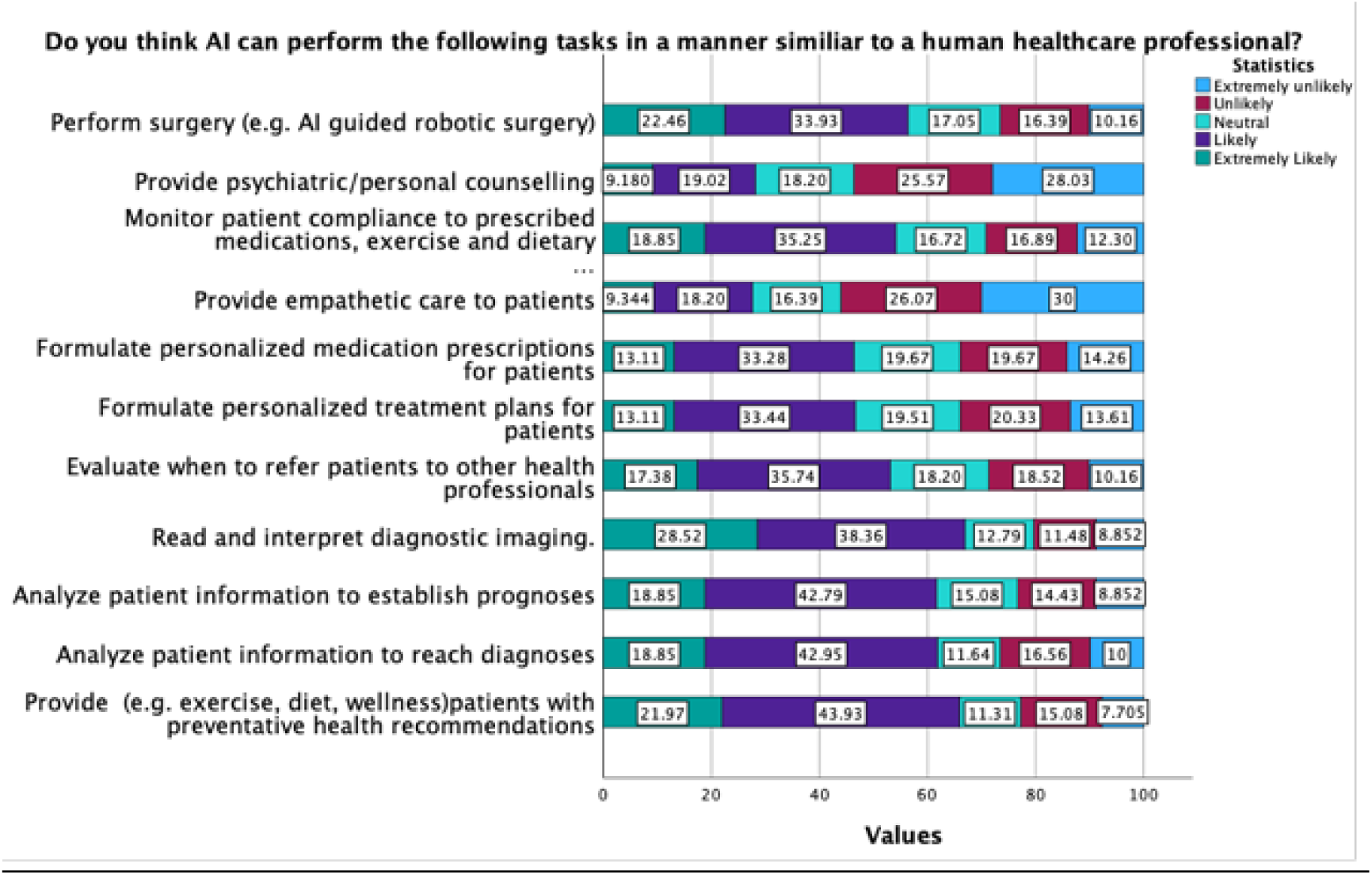
Participants were asked whether they believed AI could perform the above mentioned series of tasks at a level comparable to a human operator.

In addition, participants were asked about the potential applications of AI beyond the realm of individual health, specifically in the fields of enhancing public health and improving healthcare delivery (Fig 4).

**Fig 4.**
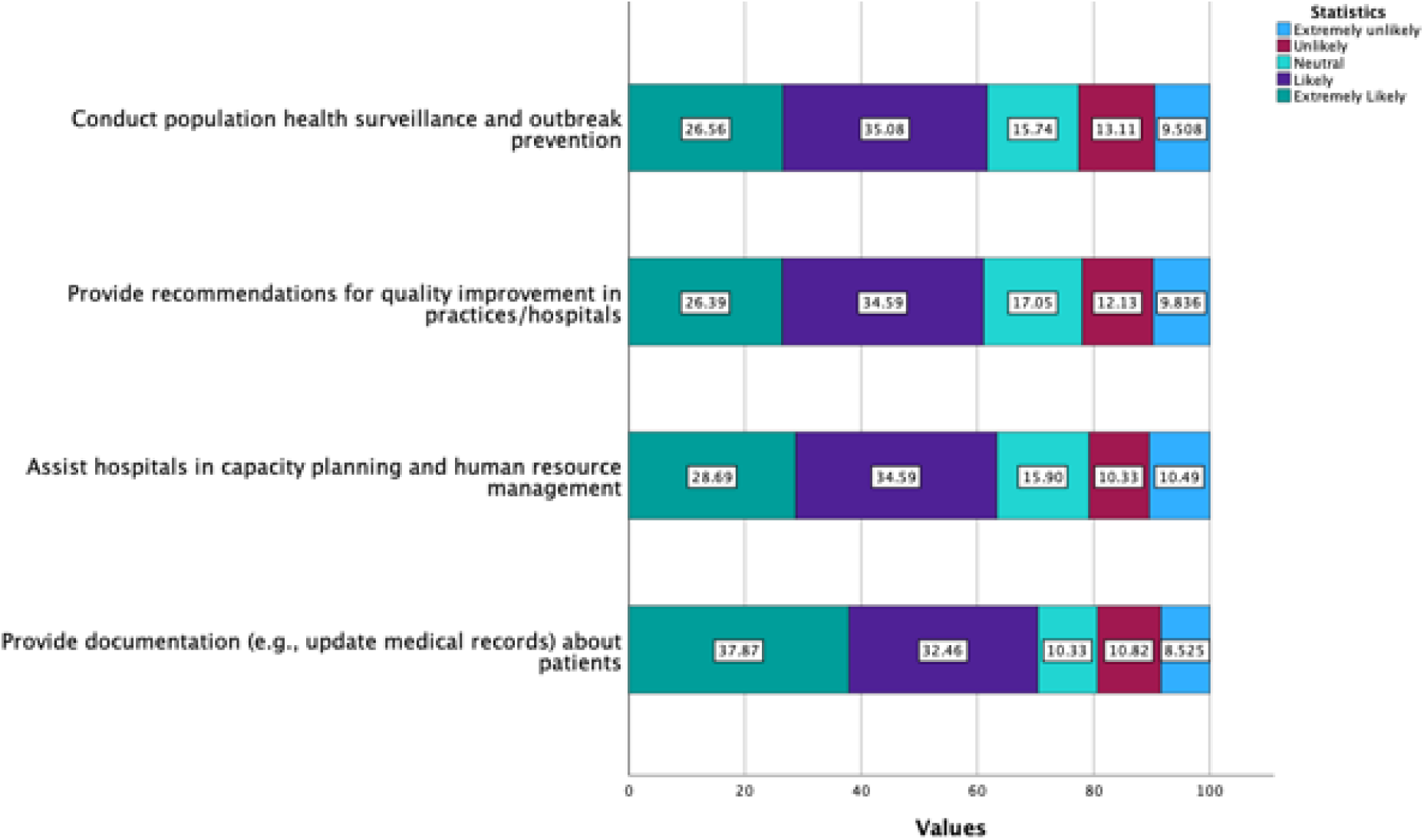
Participants were asked about the capabilities of AI in the realm of enhancing public health and healthcare delivery.

### AI’s impact on healthcare professionals and their careers/education

Participants were asked to state their levels of agreement or disagreement concerning how AI might influence their future career decisions and the significance of AI-based learning in healthcare education (Fig 5).

**Fig 5.**
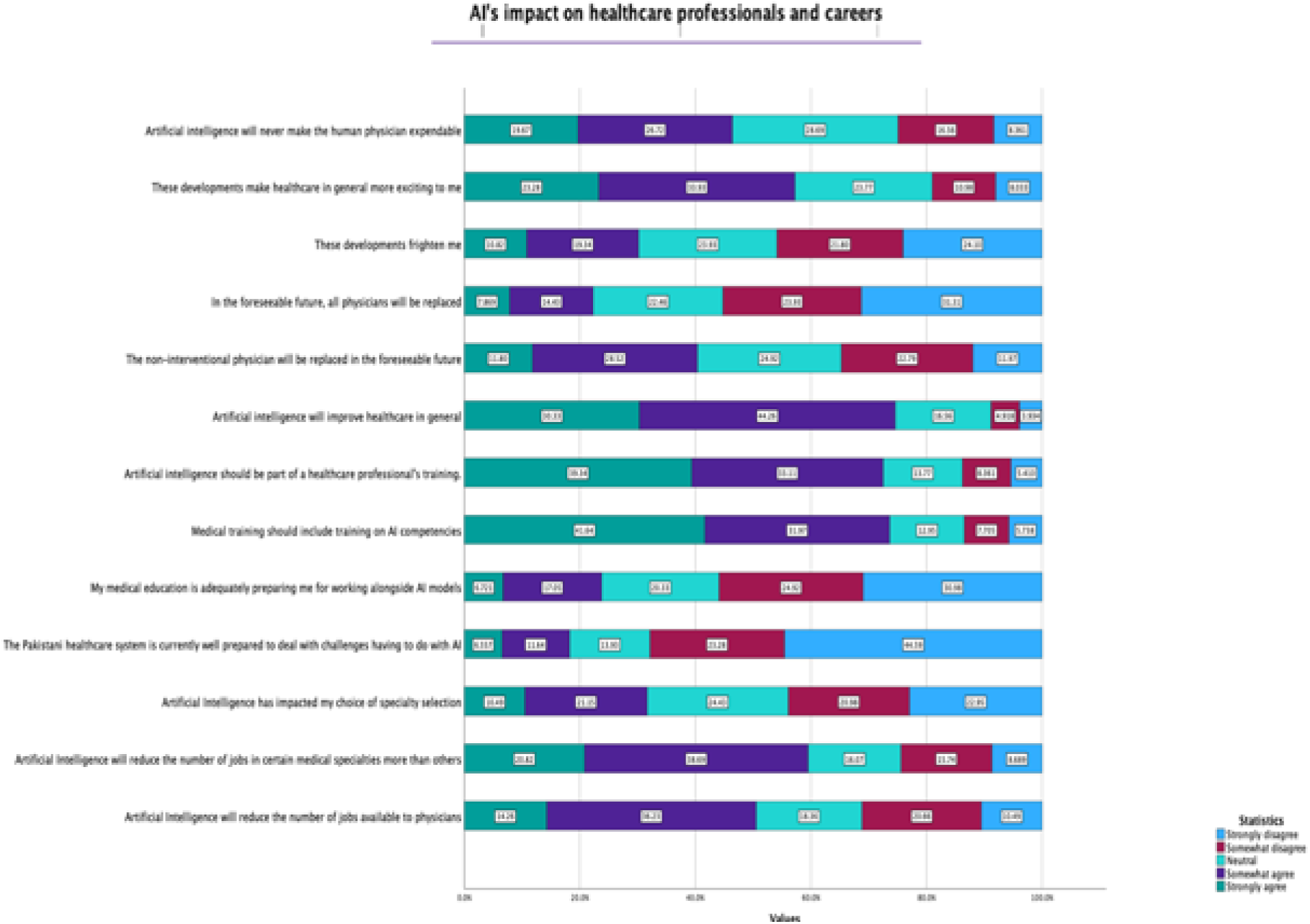
Participants were asked about their perceived impact of AI on healthcare professionals and careers.

AI’s social and ethical consequences:

Most of the participants, 70.3% (n=429), believed that AI would raise more ethical challenges in healthcare. Similarly, 73.3% (n=447) participants believed that AI would raise new social challenges, while only 12.0% (n=73) disagreed. Concerning healthcare inequity, 64.1% (n=391) of participants believed that AI would raise new health inequity issues.

The means and standard deviations of the composite scores of knowledges, attitudes and perceptions were calculated (Table 3). The mean score of knowledge of all participants was calculated to be 1.67 ± 0.342. Our findings indicate that those with higher educational qualifications (bachelor’s degrees or better) had significantly greater mean composite scores with regards to positive attitudes and perceptions about AI. We discovered no real difference with regards to knowledge, attitudes or perceptions between males and females or those with prior formal education about AI.

**Table 3.** Knowledge, attitude, and perception score of AI.

| Item | Demographic Feature | M | SD |
| --- | --- | --- | --- |
| Knowledge of AI | Female | 1.510 | 0.504 |
|  | Male | 1.489 | 0.522 |
|  | Other | 2.000 | 0.000 |
| Attitudes towards AI | Female | 2.422 | 0.838 |
|  | Male | 2.276 | 1.057 |
|  | Other | 2.133 | 0.377 |
| Perceptions of AI | Female | 2.308 | 0.588 |
|  | Male | 2.313 | 0.715 |
|  | Other | 2.500 | 0.042 |
| Knowledge of AI | Associate Ordinary Bachelor | 1.750 | 0.319 |
|  | Bachelor's degree | 1.442 | 0.542 |
|  | Below matriculation level or equivalent | 1.667 | 0.272 |
|  | Doctoral Degree | 1.588 | 0.509 |
|  | Higher Secondary School Certificate (HSSC) | 1.542 | 0.390 |
|  | Master's degree | 1.559 | 0.479 |
|  | Secondary school Certificate (SCC) - Matric | 1.333 | 0.509 |
| Attitudes<br>towards AI | Associate Ordinary Bachelor | 1.083 | 1.460 |
|  | Bachelor's degree | 2.313 | 0.943 |
|  | Below matriculation level or equivalent | 1.350 | 1.277 |
|  | Doctoral Degree | 2.506 | 0.833 |
|  | Higher Secondary School Certificate (HSSC) | 2.470 | 0.992 |
|  | Master's degree | 2.349 | 1.043 |
|  | Secondary school Certificate (SCC) - Matric | 1.324 | 1.068 |
| Perceptions of<br>AI | Associate Ordinary Bachelor | 2.279 | 1.024 |
|  | Bachelor's degree | 2.348 | 0.647 |
|  | Below matriculation level or equivalent | 1.559 | 0.933 |
|  | Doctoral Degree | 2.334 | 0.665 |
|  | Higher Secondary School Certificate (HSSC) | 2.215 | 0.574 |
|  | Master's degree | 2.268 | 0.652 |
|  | Secondary school Certificate (SCC) - Matric | 2.193 | 1.425 |
| Knowledge of<br>AI | Have had formal education in AI | 1.636 | 0.474 |
|  | Have not had formal education in AI | 1.462 | 0.519 |
| Attitudes of AI | Have had formal education in AI | 2.222 | 1.100 |
|  | Have not had formal education in AI | 2.367 | 0.934 |
| Perceptions of AI | Have had formal education in AI | 2.207 | 0.767 |
|  | Have not had formal education in AI | 2.340 | 0.631 |

## Discussion

In our survey of healthcare professionals in Pakistan, we found that although most participants lacked any formal training or experience in AI, they were generally optimistic about the capabilities of AI in healthcare. Our participants displayed good knowledge about AI and held positive attitudes and perceptions about AI’s capabilities and benefits. Specifically, they believed that AI could help establish diagnoses and prognoses based on individual factors and data, could assist hospitals in capacity planning and management and could improve healthcare in general. Most participants also believed that their medical education was not preparing them for AI technologies and competencies in healthcare. Additionally, most disagreed with the notion that the Pakistani healthcare system was well prepared to handle AI technologies. Regardless of these limitations, most participants agreed that medical professionals should receive AI training as part of their education.

Our survey also assessed participants’ general knowledge of AI and their experience with AI in education or their professional careers. Our study revealed that our study participants possessed a generally good understanding of AI and its various uses which were to a separate study conducted in Saudi Arabia. This study displayed a relatively similar mean level of knowledge of AI (2.95 ± 1.14 ) among healthcare professionals, including doctors, nurses, and technicians, regarding AI and its applications [10]. However, most of our participants did not have any formal AI training in their education or careers, which aligns with a similar study from India [11] and Nepal [12]. This is also comparable to a study from Pakistan in 2022, where more than 90% of participants had no exposure or training in AI in healthcare set-ups [13]. It is imperative for healthcare students to possess a comprehensive understanding of the underlying mechanisms through which AI technologies mediate and influence their decision-making processes. The incorporation of formalized instruction on AI concepts can serve as a springboard, enabling learners to effectively comprehend and relate to the outcomes generated by AI systems to their education and professions [14]. These benefits extend to nurses and allied health professionals as well.

More than half of our participants (59.5%, n=363) agreed that AI would substitute some healthcare professions and jobs. This finding contrasts with findings from France [15] and Syria [16] where fewer participants believed that their jobs were under threat. Despite the potential for AI to assist physicians, literature suggests that it is unlikely to replace them soon [17]. This discrepancy could be attributed to factors such as a perceived lack of confidence in technical skills, differing levels of education, economic conditions, socio-cultural factors, and may also be reflective of local industry trends. Addressing these concerns requires efforts focused on enhancing the technical proficiency of healthcare professionals, raising awareness about the collaborative potential of AI in healthcare, and the promotion of policies which support the coexistence of AI and healthcare professionals. Furthermore, participants exhibited favorable opinions regarding the potential advantages of AI in hospital systems and management. These sentiments correspond to findings in literature, which indicate that healthcare management systems powered by AI have the capacity to address various challenges, such as optimizing patient workflows in the context of a pandemic [18].

Notwithstanding, participants expressed apprehensions regarding AI’s capacity to fulfil specific roles, including the provision of psychiatric or empathetic care, as well as its potential influence on social and ethical norms and healthcare disparities. This sentiment aligns with a study from Canada which found that medical students overwhelmingly disagreed with AI’s ability to provide empathetic or psychiatric care [19]. Intriguingly, literature presents a contrasting perspective, indicating that AI chat boxes designed to respond to patient inquiries consistently generated higher quality and more empathetic responses compared to human counterparts [20][21].

Despite AI’s many applications and high demand, it is essential to consider its potential ethical and social ramifications. It is crucial for healthcare facilities, governmental and regulatory organizations to establish guidelines to tackle ethical issues and ensure accountability and responsibility [22]. Governance techniques should also be constructed to monitor the complications that may arise from AI’s integration into healthcare.

This study is the first from Pakistan to assess the knowledge, attitudes, and perceptions of healthcare professionals across the board about AI in healthcare. Previous studies from Pakistan have only surveyed doctors and medical students [8][13], while larger studies conducted internationally have either explored the latter populations or select healthcare professionals. An earlier study conducted in 2019 in Pakistan surveyed healthcare professionals, engineers, business professionals and others (e.g., researchers, data analysts etc.) but did not include medical students [23].

This survey provides novel insights for educators and stakeholders to consider when designing medical curricula or designing healthcare systems. The findings of this study will help gauge the understanding of AI that exists in the target population and potentially influence the way that medical students, doctors, nurses, and other allied health professionals perceive AI and its potential benefits. This study will also benefit current healthcare providers who can learn to actively integrate and reap the benefits of existing and developing technology.

Our research has some limitations. Firstly, we were unable to physically sample target institutions and had to rely on convenience sampling through social media, which may have introduced selection bias by reaching younger and tech savvy respondents and hence, influenced the study’s results.

### Methodology

A cross sectional study was conducted in March-April 2023 across Pakistan. The inclusion criteria included medical students, physicians, nurses, and other allied health professionals e.g., physical therapists, pharmacists and technicians from public and private medical universities, and hospitals in Pakistan that were approached via convenience sampling.

The questionnaire was prepared using Google Forms (Google, LLC) and derived its questions from 3 previously conducted studies. The questions adapted from these studies have been previously validated and have yielded good results [8][19][24]. The questionnaire was divided into 3 sub-scales namely knowledge, attitudes, and perceptions. The questions in each subsection were collected using either a 3- or 5-point Likert scale. The sub-scales were further subdivided into 5 sub-sections: (1) AI in education, (2) Basic understanding of AI, (3) AI’s capabilities in healthcare systems, (4) AI’s impact on healthcare professions and (5) AI’s social and ethical consequences. Demographic data was also collected and consisted of information included age, gender, highest qualification level, type of healthcare set-up and name, university year for the undergraduate participants and name of degree (e.g., MBBS, BScN, ASDH, PGME or others). The subscale “Knowledge of AI”, consisted of 3 questions about general understanding about AI, including knowledge of artificial AI specific terms (e.g., machine learning and deep learning), AI in daily applications and AI in surgery. Furthermore, other questions focused on whether participants had received any form of AI training during their education or careers and whether they considered themselves “technologically adept”. The subscale “Attitude towards AI” consisted of fifteen questions which included questions regarding AI’s capacity to provide preventative care, analyzing radiographical and laboratory data to make diagnoses and formulating treatment plans. The subscale “Perceptions toward artificial intelligence” consisted of ten questions which included questions on the ethical and social consequences of AI in healthcare and the impact of AI on healthcare professionals’ careers and future specializations.

A pilot was conducted on 16 participants to check for discrepancies, usability, functionality, and any further changes in the tool. The tool’s internal consistency of the assigned sub-scales was shown by Cronbach’s alpha values, ranging from 0.8 to 0.9 (Knowledge = 0.821, practice = 0.887, and Attitude = 0.930). The survey consisted of 51 prompts and the average time to complete the survey was approximately 5 minutes. The questionnaire was distributed via various social media channels to maximize the response rate for the survey, in addition to email where possible. For an expected proportion of basic AI knowledge among doctors of 27.3% [6], a sample size of 527 was required to achieve an absolute precision of ± 5% at a 99% confidence level. By considering the non-responses and incomplete data, the sample size was inflated by 15%, resulting in final sample size of 620. The sample size was estimated using Open Epi Online Sample Size calculator.

### Mean and standard deviations of knowledge, attitudes, and perception scores

The means and standard deviations of the composite scores of knowledges, attitudes and perceptions was calculated. These composite scores were then subdivided according to demographic features such as gender, education qualification and whether respondents had ever had formal education in AI (Table 3). Questions related to knowledge were of AI were graded on a 3-point Likert scale. Good knowledge was classified as a composite score of greater or equal to 1.00. Poor knowledge was classified as score less than 1.00. Questions related to attitudes and perceptions were graded on a 5-point Likert scale from 0 to 4. Mean scores greater than 2.00 signified positive attitudes and perceptions and increased agreeability. Scores less than 2.00 signified negative attitudes and perceptions and decreased agreeability.

This study received an exemption from the Aga Khan University’s Ethics Review Committee (ERC). (2023-8447-24268).

### Statistical Analysis

SPSS version (26.0) was used to process and analyze the data. The results were downloaded, imported to, and exported from, a dedicated Microsoft Excel spreadsheet. Any duplicate entries were identified and processed. Only completed responses were considered for this study. For all questions evaluated via a Likert scale, the categories were preserved and recoded numerically from 0 to 4 or from 0 to 2. Descriptive statistics included mean and medians for quantitative variables. Survey responses were summarized as frequencies and percentages.

## Discussion

In our survey of healthcare professionals in Pakistan, we found that although most participants lacked any formal training or experience in AI, they were generally optimistic about the capabilities of AI in healthcare. Our participants displayed good knowledge about AI and held positive attitudes and perceptions about AI’s capabilities and benefits. Specifically, they believed that AI could help establish diagnoses and prognoses based on individual factors and data, could assist hospitals in capacity planning and management and could improve healthcare in general. Most participants also believed that their medical education was not preparing them for AI technologies and competencies in healthcare. Additionally, most disagreed with the notion that the Pakistani healthcare system was well prepared to handle AI technologies. Regardless of these limitations, most participants agreed that medical professionals should receive AI training as part of their education.

Our survey also assessed participants’ general knowledge of AI and their experience with AI in education or their professional careers. Our study revealed that our study participants possessed a generally good understanding of AI and its various uses which were to a separate study conducted in Saudi Arabia. This study displayed a relatively similar mean level of knowledge of AI (2.95 ± 1.14 ) among healthcare professionals, including doctors, nurses, and technicians, regarding AI and its applications [12]. However, most of our participants did not have any formal AI training in their education or careers, which aligns with a similar study from India [13] and Nepal [14]. This is also comparable to a study from Pakistan in 2022, where more than 90% of participants had no exposure or training in AI in healthcare set-ups [15]. It is imperative for healthcare students to possess a comprehensive understanding of the underlying mechanisms through which AI technologies mediate and influence their decision-making processes. The incorporation of formalized instruction on AI concepts can serve as a springboard, enabling learners to effectively comprehend and relate to the outcomes generated by AI systems to their education and professions [16]. These benefits extend to nurses and allied health professionals as well.

More than half of our participants (59.5%, n=363) agreed that AI would substitute some healthcare professions and jobs. This finding contrasts with findings from France [17] and Syria [18] where fewer participants believed that their jobs were under threat. Despite the potential for AI to assist physicians, literature suggests that it is unlikely to replace them soon [19]. This discrepancy could be attributed to factors such as a perceived lack of confidence in technical skills, differing levels of education, economic conditions, socio-cultural factors, and may also be reflective of local industry trends. Addressing these concerns requires efforts focused on enhancing the technical proficiency of healthcare professionals, raising awareness about the collaborative potential of AI in healthcare, and the promotion of policies which support the coexistence of AI and healthcare professionals. Furthermore, participants exhibited favorable opinions regarding the potential advantages of AI in hospital systems and management. These sentiments correspond to findings in literature, which indicate that healthcare management systems powered by AI have the capacity to address various challenges, such as optimizing patient workflows in the context of a pandemic [20]. Notwithstanding, participants expressed apprehensions regarding AI’s capacity to fulfil specific roles, including the provision of psychiatric or empathetic care, as well as its potential influence on social and ethical norms and healthcare disparities. This sentiment aligns with a study from Canada which found that medical students overwhelmingly disagreed with AI’s ability to provide empathetic or psychiatric care [11]. Intriguingly, literature presents a contrasting perspective, indicating that AI chat boxes designed to respond to patient inquiries consistently generated higher quality and more empathetic responses compared to human counterparts [21][22].

Despite AI’s many applications and high demand, it is essential to consider its potential ethical and social ramifications. It is crucial for healthcare facilities, governmental and regulatory organizations to establish guidelines to tackle ethical issues and ensure accountability and responsibility [23]. Governance techniques should also be constructed to monitor the complications that may arise from AI’s integration into healthcare.

This study is the first from Pakistan to assess the knowledge, attitudes, and perceptions of healthcare professionals across the board about AI in healthcare. Previous studies from Pakistan have only surveyed doctors and medical students [7][8][14], while larger studies conducted internationally have either explored the latter populations or select healthcare professionals. An earlier study conducted in 2019 in Pakistan surveyed healthcare professionals, engineers, business professionals and others (e.g., researchers, data analysts etc.) but did not include medical students [24].

This survey provides novel insights for educators and stakeholders to consider when designing medical curricula or designing healthcare systems. The findings of this study will help gauge the understanding of AI that exists in the target population and potentially influence the way that medical students, doctors, nurses, and other allied health professionals perceive AI and its potential benefits. This study will also benefit current healthcare providers who can learn to actively integrate and reap the benefits of existing and developing technology.

Our research has some limitations. Firstly, we were unable to physically sample target institutions and had to rely on convenience sampling through social media, which may have introduced selection bias by reaching younger and tech savvy respondents and hence, influenced the study’s results.

## Conclusions

In conclusion, our survey provides valuable insights into healthcare professionals’ attitudes towards AI in Pakistan. Based on the survey findings we recommend an emphasis on integrating AI proficiencies into regular undergraduate education to ensure that healthcare professionals are prepared to use AI in their work. This may be in the form of training programs, seminars, and webinars related to AI, machine learning, and other AI topics These findings can inform future efforts to incorporate AI into healthcare and medical education curricula while also highlighting the need for the development of ethical and regulatory guidelines to ensure AI is used in a responsible and accountable manner.

## Ethical Considerations

Ethical approval was taken from the Ethics Review Committee at The Aga Khan University, Karachi. All procedures were performed in accordance with the ethical standards of the institutional committee. The first page of the questionnaire included the consent form which all respondents filled to proceed further. All the respondents provided consent after being instructed on the nature and purpose of the survey and were offered the possibility to withdraw at any time. Participants were also allowed to cancel the answers to their questions. All information is confidential and anonymous. Furthermore, all collected data was kept under lock and key and password protected. Access to the data was only given to the investigators. Data will only be stored for a total of 7 years in accordance with ERC guidelines.

This study is in compliance with the Good Clinical Practice (GCP) Guidelines.

## Data Availability

Data is freely available on an online Google Sheet dataset with the link attached belowhttps://docs.google.com/spreadsheets/d/1zwCGb_TCU1Cs6xm2hvLjuGpSD3jG0XsxNV3hVrkhsvw/edit?usp=sharing

https://docs.google.com/spreadsheets/d/1zwCGb_TCU1Cs6xm2hvLjuGpSD3jG0XsxNV3hVrkhsvw/edit#gid=958174516&fvid=306532044

